# Mindfulness Training and Health-Related Quality of Life in Male NCAA Division I Athletes

**DOI:** 10.1101/2023.03.09.23287051

**Authors:** Kristin Haraldsdottir, Scott Anderson, Jennifer Sanfilippo, Chad McGehee, Andrew M Watson

## Abstract

**Objective:** To examine the effects of a mindfulness intervention on mental and physical quality of life (QOL) among male collegiate athletes.

**Participants:** 327 male Division I collegiate athletes.

**Methods:** QOL was measured among 527 participants using the Veterans Rand-12 survey twice annually from 2017-2020. 101 athletes underwent 6-week mindfulness training in spring 2019. The influence of mindfulness on mental and physical QOL scores (MCS, PCS) was evaluated using linear mixed effects models with season (winter, summer), group (mindfulness, control), time (pre/post-intervention) and time*group interaction as fixed effects and individual as a random effect.

**Results:** PCS was associated with the interaction between time and group (β=1.05±0.51, p=0.04) and season (β=-0.80±0.24, p=0.001), but not time (β=-0.40±0.37, p=0.27) or group (β=0.43±0.44, p=0.33).

**Conclusion:** Mindfulness training is associated with improved physical QOL among male collegiate athletes, but did not appear to be associated with differences in mental QOL.

## Introduction

Mental health is a significant public health issue worldwide, where there is a high unmet need for interventions to provide support among this group of young adults.^1^ The college years represent a unique period of growth, where students explore identity, academic interests, interpersonal and romantic relationships, and thoughts of the future.^2^ At the same time, these years also are a period of heightened instability and uncertainty, where these new sources of growth can result in added stress, and college students are at elevated risk of mental health problems.^3^ In fact, 35% of full-time college students who were screened for six common DSM-IV mental disorders screened positive for at least one.^1^

Participation in collegiate athletics is associated with myriad quality of life-related benefits for athletes,^4^ but elite and collegiate athletes face additional, unique stressors that can contribute to compromised mental health,^5^ and interventions to support mental health are being prioritized by the National Collegiate Athletic Association and International Olympic Committee in recent years.^6,7^ Athletes experience high stress related to both sport-related factors and non-sport related factors, where thoughts of risk and fear of failure, injury, overtraining and burnout, may predispose athletes to be at greater risk of negative psychological well-being.^8^

Health-related quality of life (HRQoL) is a metric that captures information regarding aspects of life that are not generally considered physical health, but are important determinants of subjective functional and emotional well-being.^9^ Among youth athletes, it has been demonstrated that in-season injury, low sleep duration and quality, and sport specialization are associated with decreases in quality of life.^10,11^

With the recent shift to a greater emphasis on athlete mental health issues in the NCAA, the use of mindfulness among athletes has gained popularity. Mindfulness training is defined as “paying attention in a particular way: on purpose, in the present moment, and nonjudgmentally,” and is often used to provide a training vehicle for people facing mounting stress and demands to learn to relate in new ways to life’s challenges.^12^ Mindfulness training has been shown to effectively mitigate symptoms of anxiety and depression among the general population, as well as among those with cancer, and neurological disease.^13-16^ However, there is a lack of prior research evaluating the influence of mindfulness training on HRQoL among collegiate athletes. Therefore, the purpose of this study was to determine the effect of a six-week mindfulness intervention on HRQoL among male collegiate athletes during a three-year period.

## Methods

Male athletes participating in varsity sports at a Division I university completed the Veterans Rand-12 (VR-12) survey at five timepoints from 2017 to 2020. Mindfulness training was provided to the athletes by an expert with more than 10 years of experience in mindfulness teaching. The structured mindfulness training program was administered as part of regular training sessions during the spring of 2019. Each of the six, weekly half-hour sessions included procedural and declarative instruction in mindfulness practice. The in-person trainings were delivered to the participants and was made available to their respective coaching staff. The curriculum was informed by the Healthy Minds framework, and included elements of Awareness, Connection, Insight and Purpose (Figure 1).^17^

**Figure 1.**
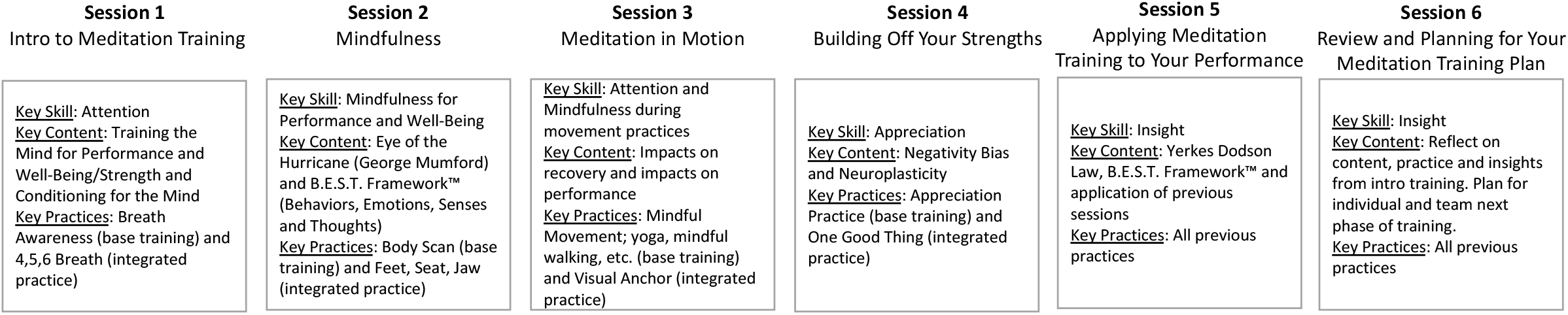
Sessions were coordinated with sport coaches and strength and conditioning coaches. Introductory sessions were required training and happened during NCAA allowed training time. Coaches were often participants in the sessions. Session length was 30 minutes. Meetings were weekly.

The VR-12 is a reliable, valid and commonly used HRQOL questionnaire that assesses the individual’s perspective on their own mental and physical health. ^18,19^ VR-12 survey responses were summarized into a physical component score (PCS) and a mental component score (MCS), where higher scores indicate better health in both domains. ^4,20^ Differences of 2 units or greater in PCS/MCS have been shown to be clinically relevant in population level data.^20,21^

Variables were initially evaluated using descriptive statistics. PCS and MCS scores were grouped by season (winter, summer) and group (mindfulness, control) for descriptive comparison between groups over time. PCS and MCS scores were then grouped by time (pre-spring 2019, post-spring 2019) for both the mindfulness and control groups. The interaction between time and mindfulness was evaluated using separate mixed effects models to predict MCS and PCS with time (pre, post), group (mindfulness, control) time of year (summer, winter) and the interaction between time and group as fixed effects, and individual as a random effect.

## Results

In total, 527 athletes participated in the study, where 101 received 6 weeks of mindfulness training in the spring of 2019. The number of survey responses at each time point is shown in Figure 2. PCS and MCS scores at each time point for each group is shown in Figure 3. The MSC and PCS scores before and after spring 2019, the time of the mindfulness intervention, are shown in Figure 4.

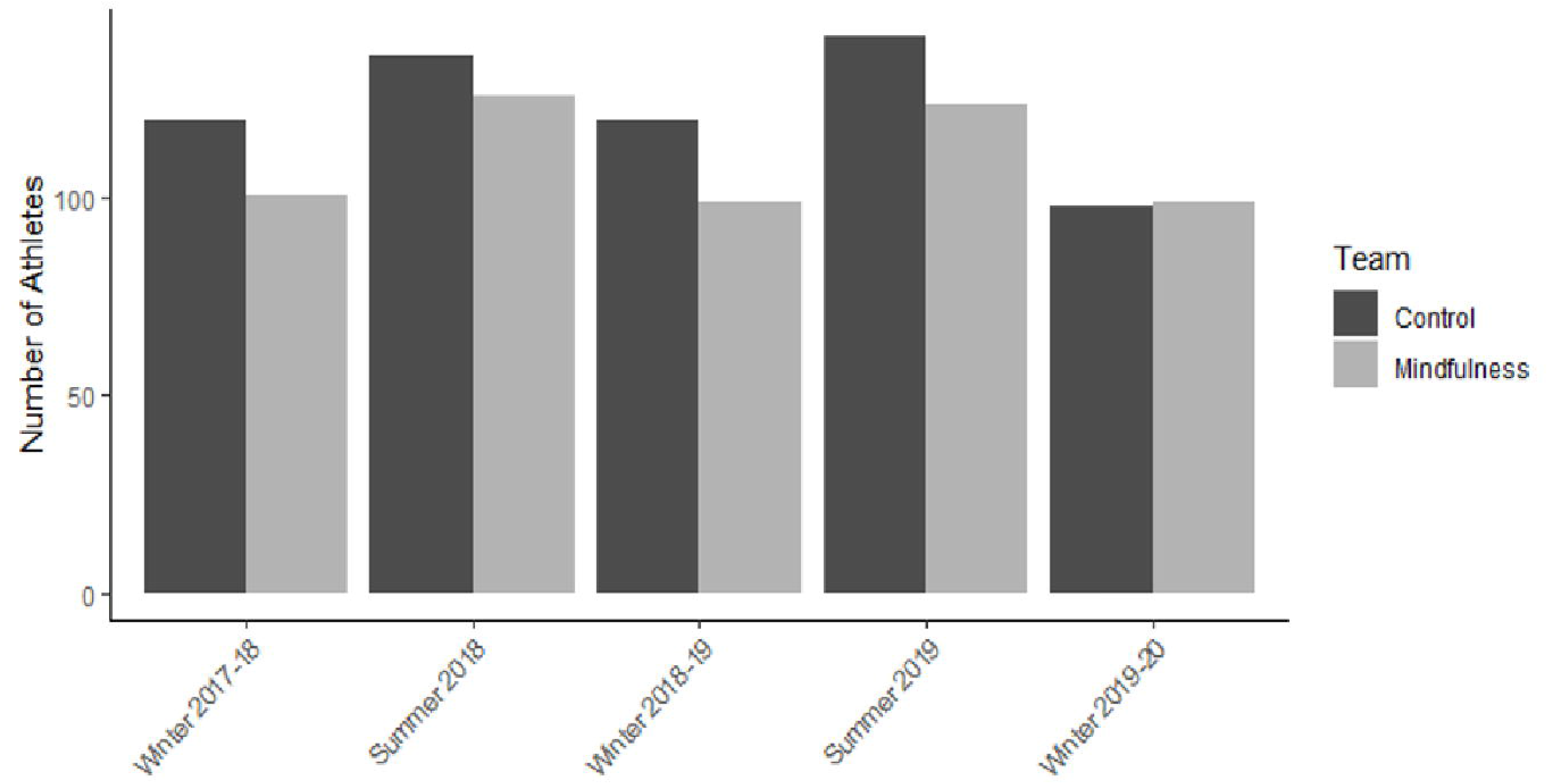

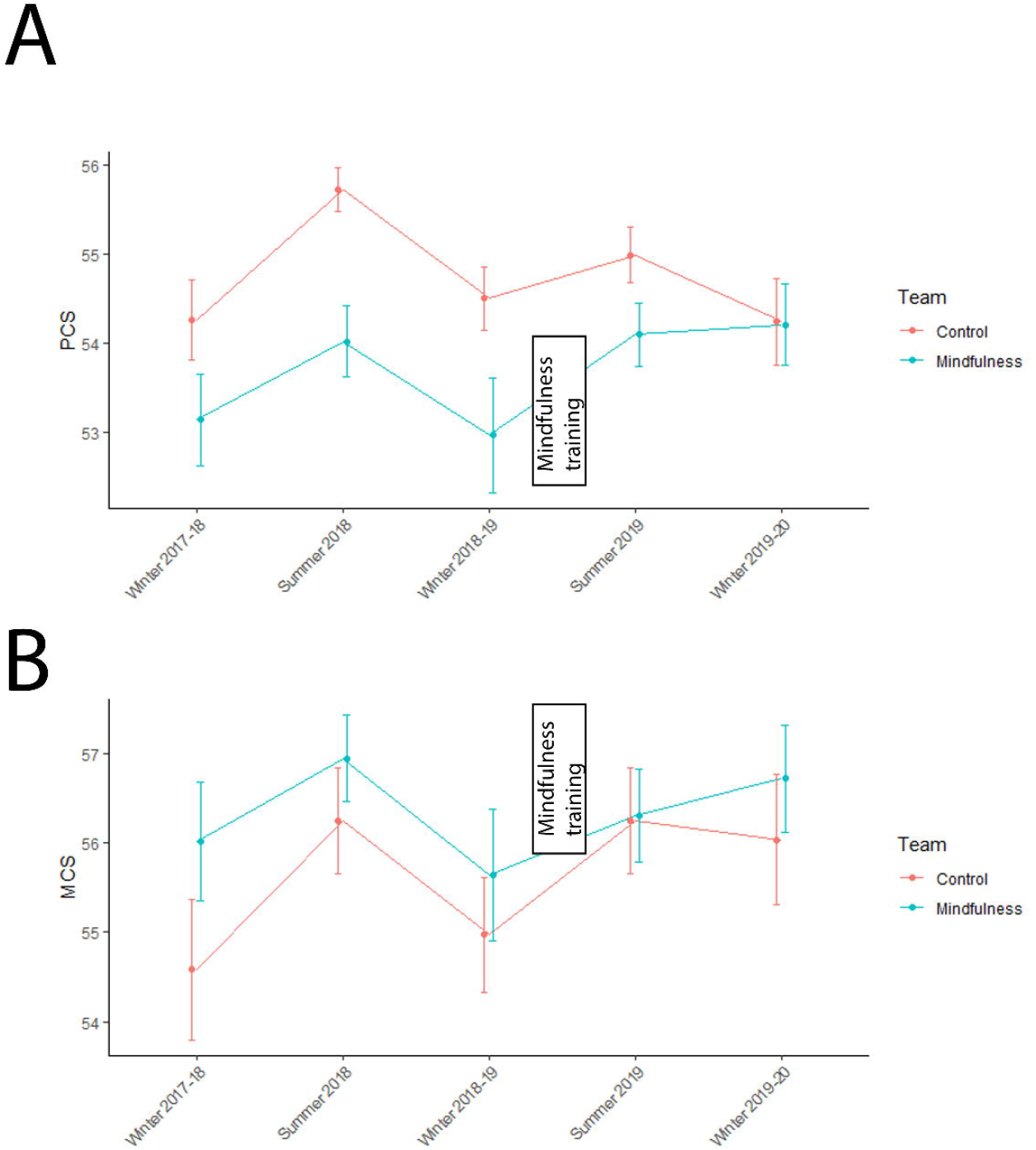

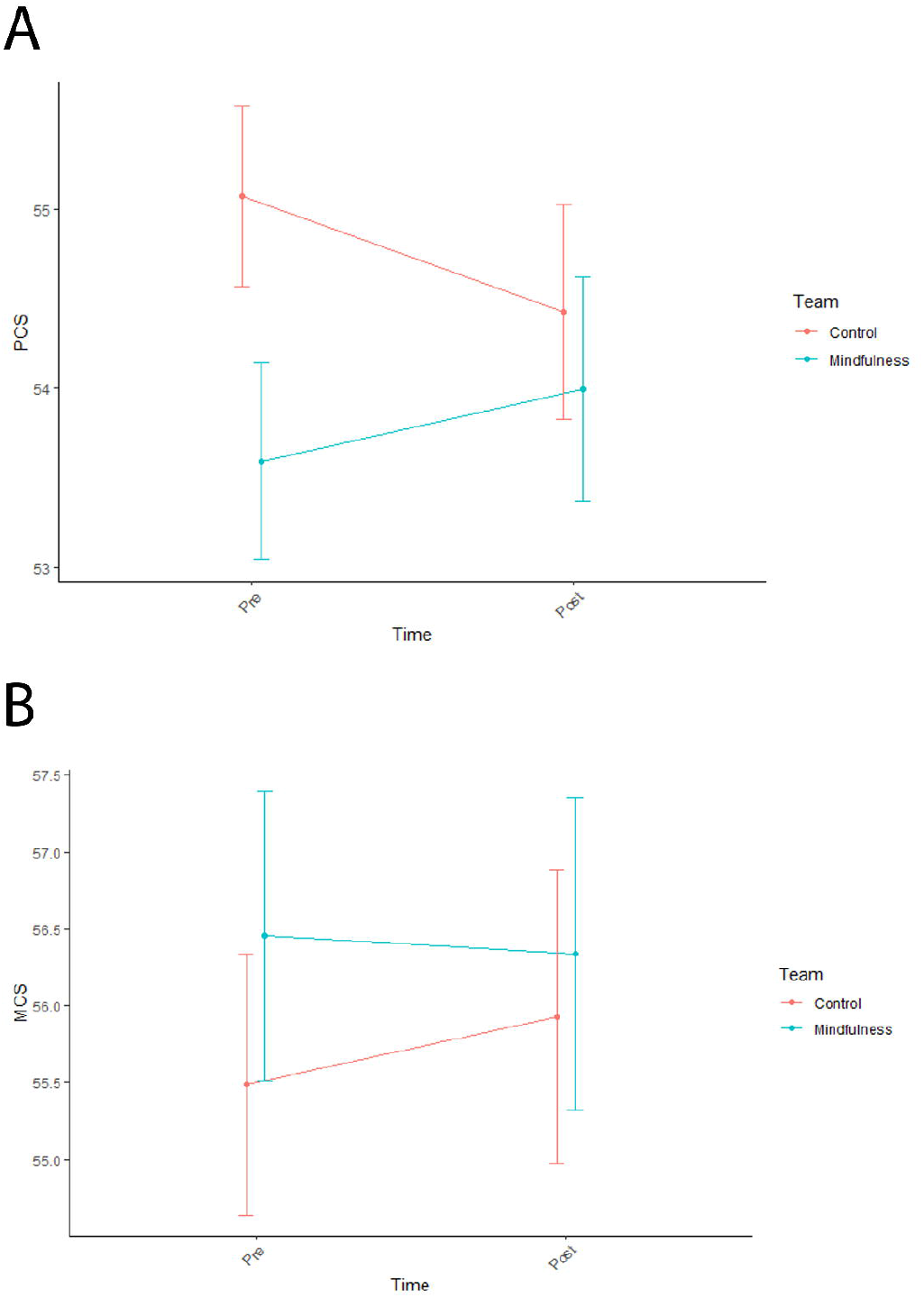

With respect to the MCS, no significant relationships were identified with respect to time (pre-mindfulness: β = 0.11 ± 0.49, p = 0.82), group (mindfulness: β = -0.41 ± 0.71, p = 0.57), or the interaction between time and group (β = -0.56 ± 0.68, p = 0.80), though there was a significant association with season, where MCS was significantly lower during the winter (β = -0.71 ± 0.31, p = 0.02). With respect to PCS, a significant interaction between time and group (β = 1.05 ± 0.51, p = 0.04) was identified. In addition, PCS was associated with season (winter: β = -0.80 ± 0.24, p = 0.001), but not with respect to time (pre-mindfulness: β = -0.40 ± 0.37, p = 0.27) or group (mindfulness: β = 0.43 ± 0.44, p = 0.33).

## Discussion

The primary aim of this study was to determine the association between a mindfulness training intervention and changes in physical and mental HRQoL among male collegiate athletes. Interestingly, while we did not find an association between mindfulness and MCS scores, we did find a significant interaction mindfulness and time with respect to PCS scores. In other words, physical QOL increased to a greater degree among the mindfulness group over time relative to the control group. Interestingly, MCS and PCS scores overall in both groups were significantly higher than values from population standards for ages 18-34 in the US ^19^, where the population average for MCS is 41.7 ± 14.3 and PCS is 40.8 ± 13.0, both nearly 15 points lower than the athlete groups in our study. This demonstrates that while mindfulness may have been associated with improvements in PCS scores, the overall HRQoL in our participant populations was high.

In comparison to male athletes who did not receive mindfulness training in the same period, we found a significant interaction between group and time, where the positive change in PCS score for the mindfulness group was significantly different than the negative change in PCS score for the control group. This difference may suggest that mindfulness training was associated with an improvement in the physical health independent of changes in the general varsity athlete population. In fact, there is recent evidence in support of the efficacy of mindfulness training and positive physical health outcomes.^22^

While we did not detect differences in the change in MCS scores between the mindfulness and control group over time, we found that MCS scores were higher in the mindfulness group during the post-season (winter) following the mindfulness training, contrary to earlier trends. While it is notable that we did not find increases in MCS scores in the mindfulness group after the mindfulness intervention, it is important to note the high baseline MCS scores in the mindfulness group, where their scores were 14.5 units higher than the population standard.^19^ Prior to the mindfulness intervention, both groups demonstrated lower MCS scores during the winter relative to the summer. Following the mindfulness intervention, however, the MCS scores in the mindfulness group did not drop during the following winter time point, while those of the control group did. Previous studies have found that athletes experience post-season stress, mental health issues and low satisfaction.^23^ The athletes in the mindfulness group all participated in a fall competition sport, and thus the winter VR-12 data collection occurred during the immediate post-season. One hypothesis to explain the steady MCS scores in the Winter 2019-20 time point in the mindfulness group, rather than a decrease, is that the mindfulness intervention may have negated the potential negative psychological effects of the post-season.

When comparing the MCS and PCS scores from the experimental and control groups with those from the general population, the values are considerably higher among the Division I student-athletes, regardless of group. In fact, the difference of an average of 12 to 14 points is well above the clinically meaningful threshold on an individual level of 6.5 units for PCS or 7.9 units for MCS.^24^ This result aligns with a previous study demonstrating that collegiate athletes have significantly higher VR-12 scores than the general population in the same age group.^4^

This study had several limitations. The investigation did not randomly assign experimental and control groups, where the mindfulness group was limited to one team that opted into a mindfulness training protocol, and thus causal relationships between group assignment not formed through randomization is not possible. Secondly, the data were collected prior to the COVID-19 pandemic, and related mental health declines associated with it.

In conclusion, we found that mindfulness training was associated with improvements in physical HRQOL among collegiate male athletes. Although we did not find a similar association with MCS overall, the mindfulness group historically exhibited a decrease in MCS during the winter (post-season) data collection, which was mitigated following mindfulness training. This suggests that mindfulness training may help athletes change their relationship to negative thoughts associated with the anxiety and challenges associated with sports participation.

## Data Availability

All data produced in the present study are available upon reasonable request to the authors.

## Acknowledgements

We acknowledge the sports medicine staff at the University of Wisconsin-Madison Division of Athletics for their commitment to the welfare of the student-athletes and for their contributions to the Badger Athletic Performance Program.

